# PSYCHIATRIC COMORBIDITY & STRUCTURAL BRAIN FEATURES IN THE ADOLESCENT BRAIN AND COGNITIVE DEVELOPMENT STUDY COHORT: A CROSS-SECTIONAL US POPULATION-BASED STUDY

**DOI:** 10.1101/2023.09.20.23295866

**Authors:** Alex Luna, Qihang Wu, Xi Zhu, Hyunnam Ryu, Rachel Marsh, Seonjoo Lee

**Affiliations:** Department of Psychiatry, New York Presbyterian Columbia University Irving Medical Center, New York, NY; New York State Psychiatric Institute, New York, NY, USA; Department of Biostatistics, Mailman School of Public health, Columbia University, New York, NY; Cognitive Neuroscience Division of the Department of Neurology and Taub Institute for Research on Alzheimer’s Disease and the Aging Brain, Vagelos College of Physicians and Surgeons, Columbia University, New York, NY; Mental Health Data Science, New York State Psychiatric Institute, New York, NY

**Keywords:** comorbidity, structural MRI, adolescent, child, network analysis, ADHD

## Abstract

**Background:** Children who develop a psychiatric disorder often also develop additional comorbid psychiatric conditions, ultimately impacting prognosis, outcomes, and treatment planning. In this cross-sectional study using the ABCD dataset, the authors set out to identify distinct comorbidity profiles using comorbidity network analysis and any associated clinical correlates of behavior and structural neuroimaging markers.

**Methods:** Structural magnetic resonance imaging and psychometric testing were obtained from 7077 eligible children between the ages of 9-10 in the ABCD dataset. Children were separated into the typically developing group and the psychiatric group based on the presence of a DSM-V diagnosis.

**Results:** Three comorbidity profiles across gender emerged using comorbidity network analysis. Girls with the ADHD – ODD (AO) comorbidity profile and sparse comorbidity profile had thicker left superior frontal gyri compared to typically developing children. Boys and girls with the ADHD – ODD comorbidity profile had significantly higher externalizing scores compared to typically developing children. The ADHD-OCD-Specific Phobia (AOS) profile among boys had significantly higher internalizing scores, while the AO profile had significantly higher internalizing scores for girls. The AOS profile for boys and the AO profile for girls had significantly higher total problem scores compared to typically developing children.

**Conclusion:** Comorbidity network analysis successfully identified comorbidity profiles associated with unique neurobiological markers and behavioral correlates and is a feasible technique for the investigation of comorbid psychiatric conditions.

## Introduction

Approximately 50% of all adolescents will develop a psychiatric disorder throughout their lifetime and 27.9% will develop two or more comorbid disorders.(1, 2) Studies have demonstrated that comorbidity occurs in a complex, heterogeneous manner. For example, 73% of children with major depressive disorder (MDD) also develop comorbid anxiety.(3) Among children with Attention Deficit Hyperactivity Disorder (ADHD), 60% - 100% develop one or more comorbid disorders, such as generalized anxiety disorder (GAD), conduct disorder (CD), and obsessive-compulsive disorder (OCD).(4-7) Children diagnosed with specific phobia are 2.5 times more likely to have GAD and 8 times as likely to develop oppositional defiant disorder (ODD).(8)

Comorbidity can severely impact outcomes for youth. In patients with both ADHD and OCD, symptom persistence, symptom severity, and overall prognosis are worse in comparison to those with OCD alone.(9, 10) Furthermore, comorbidity impacts the effectiveness of treatment, as shown in a study of children with specific phobia whose comorbidity with ADHD predicted poorer immediate and long-term treatment outcomes.^(11)^ The presence of multiple comorbid psychiatric disorders in children and adolescents is also associated with higher rates of adverse outcomes, such as social impairment, and suicide attempts.(12, 13)

Cortical thickness, derived from T1 imaging, plays an important role in several psychiatric disorders.(14) For example, decreased cortical thickness of frontotemporal, parietal, and occipital cortices is observed in schizophrenia,(15) while increased cortical thickness is observed across numerous structures of the default mode network in depression.(16) However, such studies are often limited to patients with few to no comorbidities, are seldom done with children, or tend to solely focus on comorbidities with ADHD.(17, 18) Though such studies suggest possible overlapping biomarkers in psychiatric comorbidity, limitations such as non-standardized image collection, lack of internal and external validation, and small sample sizes restrict their generalizability.(19) Furthermore, group-level analysis limits the assessment of complex comorbidity patterns involving multiple diagnoses.

Large-scale population-based datasets like the Adolescent Brain Cognitive Development (ABCD) allow for studying comorbidity biomarkers with greater generalizability. However, the high-dimensional interplay of comorbid psychiatric disorders poses significant challenges, especially in children who may be diagnosed with up to 7 comorbid disorders at any one time.(20) Network analysis has been used in adult comorbidity research to identify associations between psychiatric symptoms and physical symptoms across diagnoses and to identify the most salient symptoms occurring across anxiety and depression, allowing for a nuanced understanding of this comorbidity profile. (21, 22) Yet, network analysis remains unexplored in youth psychiatric disorders and across multiple comorbid diagnoses. Combining network analysis with clustering methods and neuroimaging has the potential to identify prominent comorbidity profiles and associated neuroimaging features in youth, an area yet to be explored.

In this data-driven cross-sectional study, we applied network analysis to psychiatric diagnoses in a large, heterogeneous sample of youth to identify several distinct comorbidity profiles. We hypothesized that specific neuroimaging biomarkers unique to each comorbidity profile would emerge and associate with a unique behavioral profile.

## Methods and Materials

### 2.1 ABCD Dataset

The Adolescent Brain Cognitive Development (ABCD) is a large-scale study of 11,878 youth aged 9–10 years recruited from 21 research sites across the United States. Participants were excluded if they were not proficient in English, or if parents were not fluent in English or Spanish; had a severe sensory, intellectual, medical, or neurological condition that would make their data invalid or limit their ability to follow the study protocols; or had contraindications to undergoing MRI.(23) Data was collected from baseline visits between September 1, 2016 and November 15, 2018 and analyses were conducted on the ABCD Study 3.0 data release.(24) A single IRB is maintained by the University of California San Diego Human Research Protections Program (160091). All parents provided written informed consent and all children provided assent.

All participants across the sites underwent MRI imaging using scanners from GE Healthcare (Waukesha, Wisconsin), Siemens Healthcare (Erlangen, Germany), or Philips Healthcare (Andover, Massachusetts). More detailed descriptions of the MRI acquisition protocol are found elsewhere.(24) In brief, structural MRI in the ABCD cohort was conducted using 3T MRI scanners, yielding high resolution T1 weighted images (1mm isotropic). Images were processed using a minimally processed pipeline and freesurfer v7.0.1 (https://surfer.nmr.mgh.harvard.edu). Cortical thickness values were extracted using the Desikan-Killiany-Tourville (DKT) atlas, and subcortical volumes were calculated using the ASEG atlas.(25, 26) In total 62 bilateral cortical thickness measures and 21 subcortical volume measures (Thalamus, Caudate, Putamen, Pallidum, Hippocampus, Amygdala, Accumbens area, Ventral DC, CC Posterior, CC Mid Posterior, CC Central, CC Mid Anterior, and CC Anterior) were used for further analysis.(27)

In this study, we used a subset of past and present DSM-V diagnoses from the Computerized Kiddie-Structured Assessment for Affective Disorders and Schizophrenia for DSM-V (KSADS-COMP.) The KSADS-COMP was the primary clinical measure utilized for diagnostic assessment due to its availability in the ABCD dataset, consistency with the DSM-V, use in research settings, and favorable validity data. Participants with no responses and variables with high rates of missing values or no “Yes” responses were removed, leaving a total of 11,716 participants. Among them, 4,499 TD children were selected based on the absence of any past or present DSM-V diagnoses. Similarly, for the psychiatric group, 2,578 children were selected based on having at least one current DSM-V diagnosis. Participants with bipolar disorder were excluded from this analysis due to concerns about a falsely elevated prevalence in the ABCD dataset at baseline.(28) More detailed information about the data cleaning process can be found in eFigure 1 in the Supplement.

Individual participants in the ABCD dataset underwent a battery of psychological tests, including the Child Behavior Checklist (CBCL) and the NIH-Toolbox for neurocognitive testing. (29-32) Demographic information was collected from caregivers.

### 2.2 Statistical Analysis

Data cleaning and all statistical analyses were performed in R and were stratified by sex given well-documented reports of gender differences in psychiatric disorders.(33) For further detail regarding analyses, including the participant selection pipeline (eFigure 1), and comorbidity network analysis employed for identification of comorbidity profiles (eFigure 2), please refer to the provided supplemental material. In brief, among 7077 participants, frequencies of pairs of co-occurring diagnoses were used to determine comorbidity profiles using a combination of Bayesian Information Criteria (BIC) and latent class analysis (LCA). Once comorbidity profiles were established, linear mixed effect modeling was used to compare subcortical and cortical measures between each group with site as random intercepts. Behavioral profile analysis was conducted using ANOVAs to examine differences in CBCL scores and NIH-Toolbox metrics across comorbidity profiles and sex. Post-hoc pair-wise group comparisons were performed using the Benjamini-Hochberg adjustment. Of note, for only the structural MRI analysis, only 6676 participants (4269 in the TD group and 2407 in the psychiatric group) were included as 401 participants of the 7077 participants initially selected did not have high quality MRI data available.

## Results

### 3.1 baseline demographics

This study included 7,077 participants from the ABCD cohort. The sample was composed of 3,476 total females (49.1%) and 3,601 males (50.8%). The average age for boys was 9.93 years (SD = 0.62 years), 9.89 years (SD = 0.62) for girls (Tables 1 & 2). Regarding ethnic composition, 75% of boys and 71% of girls identified as white, 21% of boys and 24% of girls identified as black, 21% of boys and girls identified as Hispanic, and 4.7% of boys and 4.9% of girls identified as other. Of note, some individuals were noted to have selected multiple ethnic categories.

**Table 1.** Baseline demographics and psychological metrics for boys.

| Characteristics | Participants, No.<br>(%) |  |  |  |  | *P* value<sup>b</sup> |
| --- | --- | --- | --- | --- | --- | --- |
|  | Overall<br>(N = 3601) | TD<br>(n = 2113)<sup>a</sup> | AOS<br>(n = 74) | AO<br>(n = 214) | SPA<br>(n = 1200) |  |
| Demographic characteristics |  |  |  |  |  |  |
| Age, mean (SD), y | 9.93 (0.62) | 9.95 (0.62) | 10.04<br>(0.59) | 9.92<br>(0.62) | 9.88 (0.63) | 0.01 |
| Race |  |  |  |  |  |  |
| White | 2702 (75) | 1585 (75) | 52 (70) | 169 (79) | 896 (75) | 0.4 |
| Black | 752 (21) | 406 (19) | 26 (35) | 50 (23) | 270 (22) | 0.002 |
| Other race | 168 (4.7) | 82 (3.9) | 6 (8.1) | 13 (6.1) | 67 (5.6) | 0.04 |
| Ethnicity |  |  |  |  |  |  |
| Hispanic | 733 (21) | 463 (22) | 14 (19) | 24 (11) | 232 (20) | 0.002 |
| Not documented, No. | 50 | 22 | 2 | 4 | 22 |  |
| Parental education level |  |  |  |  |  |  |
| Below college | 630 (18) | 402 (19) | 15 (20) | 30 (14) | 183 (15) | 0.02 |
| At least some college | 2963 (82) | 1708 (81) | 59 (80) | 183 (86) | 1013 (85) |  |
| Not documented, No. | 8 | 3 | 0 | 1 | 4 |  |
| Household income |  |  |  |  |  |  |
| <\$50,000 | 938 (29) | 496 (26) | 31 (46) | 49 (26) | 362 (33) | <0.001 |
| ≥\$50,000 | 2346 (71) | 1420 (74) | 36 (54) | 140 (74) | 750 (67) | |
| Not documented, No. | 317 | 197 | 7 | 25 | 88 |  |
| Parental substance use I<sup>c</sup> |  |  |  |  |  |  |
| Tobacco | 466 (13) | 208 (10.0) | 15 (21) | 42 (20) | 201 (17) | <0.001 |
| Alcohol | 826 (24) | 414 (21) | 17 (24) | 63 (33) | 332 (30) | <0.001 |
| Others | 219 (6.1) | 82 (3.9) | 11 (15) | 22 (10) | 104 (8.7) | <0.001 |
| **Parental substance use II<sup>d</sup>** |  |  |  |  |  |  |
| **Tobacco** | 171 (4.8) | 64 (3.1) | 8 (11) | 18 (8.8) | 81 (7.0) | <0.001 |
| **Others** | 72 (2.0) | 19 (0.9) | 4 (5.4) | 9 (4.2) | 40 (3.3) | <0.001 |
| **Born prematurely (<37 weeks)** | 477 (13) | 282 (13) | 11 (15) | 28 (13) | 156 (13) | 0.9 |
| **Not documented, No.** | 43 | 14 | 3 | 1 | 25 |  |
| **Psychological metrics** |  |  |  |  |  |  |
| **CBCL** |  |  |  |  |  |  |
| **External** | 46 (11) | 41 (8) | 59 (10) | 64 (9) | 51 (10) | <0.001 |
| **Internal** | 49 (11) | 44 (9) | 66 (7) | 57 (10) | 54 (10) | <0.001 |
| **TotProb** | 46 (12) | 39 (9) | 66 (7) | 63 (8) | 53 (10) | <0.001 |
| **NIH ToolBox** |  |  |  |  |  |  |
| **Crystallized** | 51 (11) | 52 (11) | 49 (11) | 49 (11) | 50 (11) | <0.001 |
| **Not documented, No.** | 294 | 166 | 4 | 21 | 103 |  |
| **Cognition fluid** | 46 (11) | 46 (11) | 43 (12) | 44 (11) | 44 (11) | <0.001 |
| **Not documented, No.** | 307 | 172 | 5 | 22 | 108 |  |
| **Cognition total** | 48 (11) | 49 (11) | 45 (12) | 45 (11) | 46 (11) | <0.001 |
| **Not documented, No.** | 307 | 172 | 5 | 22 | 108 |  |

Abbreviations: **TD**, Typically Developing; **AOS**, ADHD+OCD+SPH; **AO**, ADHD+ODD; **SPA**, Sparse Clusters; **CBCL**, Child Behavior Checklist; **TotProb**, Total Problems.
<sup>a</sup> Removed one boy due to missing CBCL scores.
<sup>b</sup> One-way ANOVA for continuous variables; Pearson's Chi-square test for categorical variables.
<sup>c</sup> Substance use prior to pregnancy (missing values are not displayed).
<sup>d</sup> Substance use after knowing of pregnancy (missing values are not displayed).

**Table 2.** Baseline demographics and psychological metrics for girls.

| Characteristics | Participants, No. (%) |  |  |  |  | P value<sup>b</sup> |
| --- | --- | --- | --- | --- | --- | --- |
|  | Overall<br>(N = 3476) | TD<br>(n = 2386)<sup>a</sup> | SPH<br>(n = 425) | AO<br>(n = 88) | SPA<br>(n = 577) |  |
| Demographic characteristics |  |  |  |  |  |  |
| Age, mean (SD), y | 9.89 (0.62) | 9.89 (0.62) | 9.90 (0.63) | 9.94 (0.62) | 9.89 (0.62) | 0.8 |
| Race |  |  |  |  |  |  |
| White | 2485 (71) | 1675 (70) | 295 (69) | 63 (72) | 452 (78) | 0.001 |
| Black | 826 (24) | 544 (23) | 132 (31) | 24 (27) | 126 (22) | 0.001 |
| Other race | 172 (4.9) | 100 (4.2) | 35 (8.2) | 3 (3.4) | 34 (5.9) | 0.003 |
| Ethnicity |  |  |  |  |  |  |
| Hispanic | 734 (21) | 526 (22) | 95 (23) | 8 (9.1) | 105 (19) | 0.007 |
| Not documented, No. | 43 | 28 | 4 | 0 | 11 |  |
| Parental education level |  |  |  |  |  |  |
| Below college | 626 (18) | 422 (18) | 99 (23) | 18 (20) | 87 (15) | 0.008 |
| At least some college | 2848 (82) | 1962 (82) | 326 (77) | 70 (80) | 490 (85) |  |
| Not documented, No. | 2 | 2 | 0 | 0 | 0 |  |
| Household income |  |  |  |  |  |  |
| <\$50,000 | 984 (31) | 639 (29) | 155 (40) | 32 (41) | 158 (30) | <0.001 |
| ≥\$50,000 | 2181 (69) | 1530 (71) | 236 (60) | 47 (59) | 368 (70) | |
| Not documented, No. | 311 | 217 | 34 | 9 | 51 |  |
| Parental substance use I<sup>c</sup> |  |  |  |  |  |  |
| Tobacco | 478 (14) | 260 (11) | 88 (21) | 23 (28) | 107 (19) | <0.001 |
| Alcohol | 828 (25) | 549 (24) | 98 (25) | 22 (27) | 159 (30) | .05 |
| Others | 246 (7.1) | 127 (5.3) | 38 (8.9) | 16 (18) | 65 (11) | <0.001 |
| Parental substance use II<sup>d</sup> |  |  |  |  |  |  |
| **Tobacco** | 180 (5.3) | 85 (3.6) | 41 (9.9) | 14 (17) | 40 (7.3) | <0.001 |
| **Others** | 103 (3.0) | 46 (1.9) | 13 (3.1) | 9 (10) | 35 (6.1) | <0.001 |
| **Born prematurely (<37 weeks)** | 455 (13) | 319 (14) | 56 (13) | 11 (13) | 69 (12) | 0.8 |
| **Not documented, No.** | 53 | 31 | 4 | 5 | 13 |  |
| **Psychological metrics** |  |  |  |  |  |  |
| **CBCL** |  |  |  |  |  |  |
| **External** | 44 (10) | 41 (8) | 48 (10) | 64 (9) | 53 (10) | <0.001 |
| **Internal** | 46 (11) | 43 (8) | 52 (11) | 61 (11) | 55 (10) | <0.001 |
| **TotProb** | 44 (11) | 39 (9) | 50 (10) | 64 (8) | 55 (9) | <0.001 |
| **NIH ToolBox** |  |  |  |  |  |  |
| **Crystallized** | 51 (12) | 51 (12) | 50 (12) | 49 (13) | 50 (11) | 0.01 |
| **Not documented, No.** | 283 | 201 | 24 | 4 | 54 |  |
| **Cognition fluid** | 46 (11) | 47 (11) | 46 (12) | 44 (13) | 44 (11) | <0.001 |
| **Not documented, No.** | 300 | 209 | 25 | 5 | 61 |  |
| **Cognition total** | 48 (11) | 49 (11) | 47 (12) | 45 (14) | 46 (11) | <0.001 |
| **Not documented, No.** | 304 | 212 | 26 | 5 | 61 |  |

Abbreviations: **TD**, Typically Developing; **SPH**, Specific Phobia; **AO**, ADHD+ODD; **SPA**, Sparse Clusters; **CBCL**, Child Behavior Checklist; **TotProb**, Total Problems.
<sup>a</sup> Removed three girls due to missing CBCL scores.
<sup>b</sup> One-way ANOVA for continuous variables; Pearson's Chi-squared test for categorical variables.
<sup>c</sup> Substance use prior to pregnancy (missing values are not displayed).
<sup>d</sup> Substance use after during pregnancy (missing values are not displayed).

Baseline demographics were collected across all children with psychiatric diagnoses and compared to typically developing children (Tables 1 & 2). Significant differences were observed in household income, parental substance use, parental education level, ethnicity, and race when compared to typically developing children. No significant differences were observed in relation to prematurity (Tables 1 & 2).

### 3.2 Psychiatric Comorbidity Cluster Analysis

Cluster analysis yielded 3 distinct profiles for both boys and girls. Each profile was characterized by the most prevalent comorbidities within each group (eFigure 3). Comorbid diagnoses were deemed predominant if over 50% of the participants within a profile had comorbidity across these diagnoses (Table 1).

Among boys, one profile comprised 74 participants with predominant comorbidity across ADHD, OCD, and specific phobia (AOS). The second profile consisted of 214 participants, primarily presenting comorbidity between ADHD and ODD (AO). The third profile, referred to as the sparse comorbidity profile (SPA), included 1,200 individuals and did not have any predominant comorbid diagnoses.

Among girls, the first profile comprised 425 participants, all diagnosed with specific phobia, but without any comorbid diagnoses meeting the 50% threshold (SPH). The second profile featured comorbid ADHD and ODD (AO) and consisted of 88 participants. The final profile with 577 participants, did not have any predominant comorbid diagnoses, resulting in a sparse comorbidity (SPA) designation.

Table 1 presents the specific prevalence values for the top 10 comorbid diagnoses in each profile across sex, while the prevalence values for each individual diagnosis across each profile can be found in Table 3. In the boys’ AOS profile, 63.5% had comorbid ADHD and SPH, 62.2% had comorbid ADHD and OCD, and 52.7% had comorbid OCD and SPH. Notable comorbid diagnoses with over 30% prevalence included ADHD and ODD (43.8%), OCD and ODD (35.1%) SPH and ODD (31.1%), and GAD and ADHD (36.5%). In the boys’ AO profile, 73.8% had comorbid ADHD and ODD, 33.6% had comorbid ADHD and conduct disorder (CD), and 30.8% had comorbid ODD and CD. No comorbid diagnoses exceeded 10% prevalence in the SPA profile.

**Table 3.** Sex stratified prevalence values for each DSM-V diagnosis across comorbidity profiles.

| Diagnoses<sup>a</sup> | Prevalence, % |  |  |  |  |  |  |  |
| --- | --- | --- | --- | --- | --- | --- | --- | --- |
|  | Boys |  |  |  | Girls |  |  |  |
|  | Overall<br>(N = 1488) | AOS<br>(n = 74) | AO<br>(n = 214) | SPA<br>(n = 1200) | Overall<br>(N = 1090) | SPH<br>(n = 425) | AO<br>(n = 88) | SPA<br>(n = 577) |
| **Attention Deficit Hyperactivity Disorder (ADHD)** | 43.35 | 83.78 | 88.32 | 32.83 | 21.16 | 8.71 | 70.46 | 34.14 |
| **Anorexia Nervosa (AN)** | 0.07 | 1.35 | 0.00 | 0.00 | 0.00 | 0.00 | 0.00 | 0.00 |
| **Binge-eating Disorder (BED)** | 2.29 | 0.00 | 2.80 | 2.33 | 2.84 | 1.88 | 5.68 | 3.12 |
| **Bulimia Nervosa (BN)** | 0.07 | 0.00 | 0.00 | 0.08 | 0.28 | 0.00 | 0.00 | 0.52 |
| **Conduct Disorder (CD)** | 12.23 | 12.16 | 44.86 | 6.42 | 7.98 | 0.00 | 42.05 | 8.67 |
| **Generalized Anxiety Disorder (GAD)** | 3.43 | 43.24 | 0.00 | 1.58 | 4.59 | 2.12 | 26.14 | 3.12 |
| **Major Depressive Disorder (MDD)** | 0.94 | 12.16 | 0.00 | 0.42 | 0.83 | 0.00 | 7.96 | 0.35 |
| **Obsessive Compulsive Disorder (OCD)** | 30.24 | 74.32 | 19.16 | 29.50 | 27.62 | 9.65 | 46.59 | 37.96 |
| **Oppositional Defiant Disorder (ODD)** | 23.93 | 44.60 | 85.05 | 11.75 | 19.63 | 4.47 | 65.91 | 23.74 |
| **Panic Disorder (PD)** | 0.07 | 1.35 | 0.00 | 0.00 | 0.18 | 0.00 | 1.14 | 0.17 |
| **Post-traumatic Stress Disorder (PTSD)** | 0.40 | 6.76 | 0.47 | 0.00 | 0.18 | 0.00 | 2.27 | 0.00 |
| **Schizophrenia (SCZ)** | 1.08 | 10.81 | 0.00 | 0.67 | 0.73 | 0.47 | 0.00 | 1.04 |
| **Separation Anxiety Disorder (SEP)** | 1.41 | 21.62 | 0.00 | 0.42 | 1.01 | 0.00 | 10.23 | 0.35 |
| **Social Anxiety Disorder (SOC)** | 2.89 | 5.41 | 6.54 | 2.08 | 4.86 | 4.94 | 4.55 | 4.85 |
| **Specific Phobia (SPH)** | 31.65 | 72.97 | 15.42 | 32.00 | 43.21 | 100.00 | 52.27 | 0.00 |

Abbreviations: **AOS**, ADHD+OCD+SPH; **AO**, ADHD+ODD; **SPH**, Specific Phobia; **SPA**, Sparse Clusters.
<sup>a</sup> 15 Current DSM-V diagnoses (abbreviation).

Among girls in the AO profile, 52.3% had comorbid ADHD and ODD, while 34.1% had comorbid ADHD and OCD. Neither the SPH nor the SPA profiles reached a prevalence higher than 10%. A Chi-square analysis was conducted on all three profiles by gender, revealing a significant groupwise difference in diagnostic prevalence for both boys and girls (eTable 2).

### 3.3 Cortical and subcortical imaging measures

Cortical thickness and subcortical volume were compared between TD children and those with the identified comorbidity profiles. No significant differences in those measures were detected in boys. In contrast, a significant difference in bilateral superior frontal cortices was detected in girls across the three profiles. Post-hoc analysis revealed thicker bilateral superior frontal cortices for the AO and SPA profiles compared to the TD group among girls (AO: Estimate = 0.03, 95% CI 0.00 - 0.05, P = 0.05, SPA: Estimate = 0.02, 95% CI 0.01 - 0.03, P = 0.002, Figure 1). No significant differences were identified between the SPH profile and the TD group, nor between the AO profile and the SPA profile in girls.

**Figure 1.**
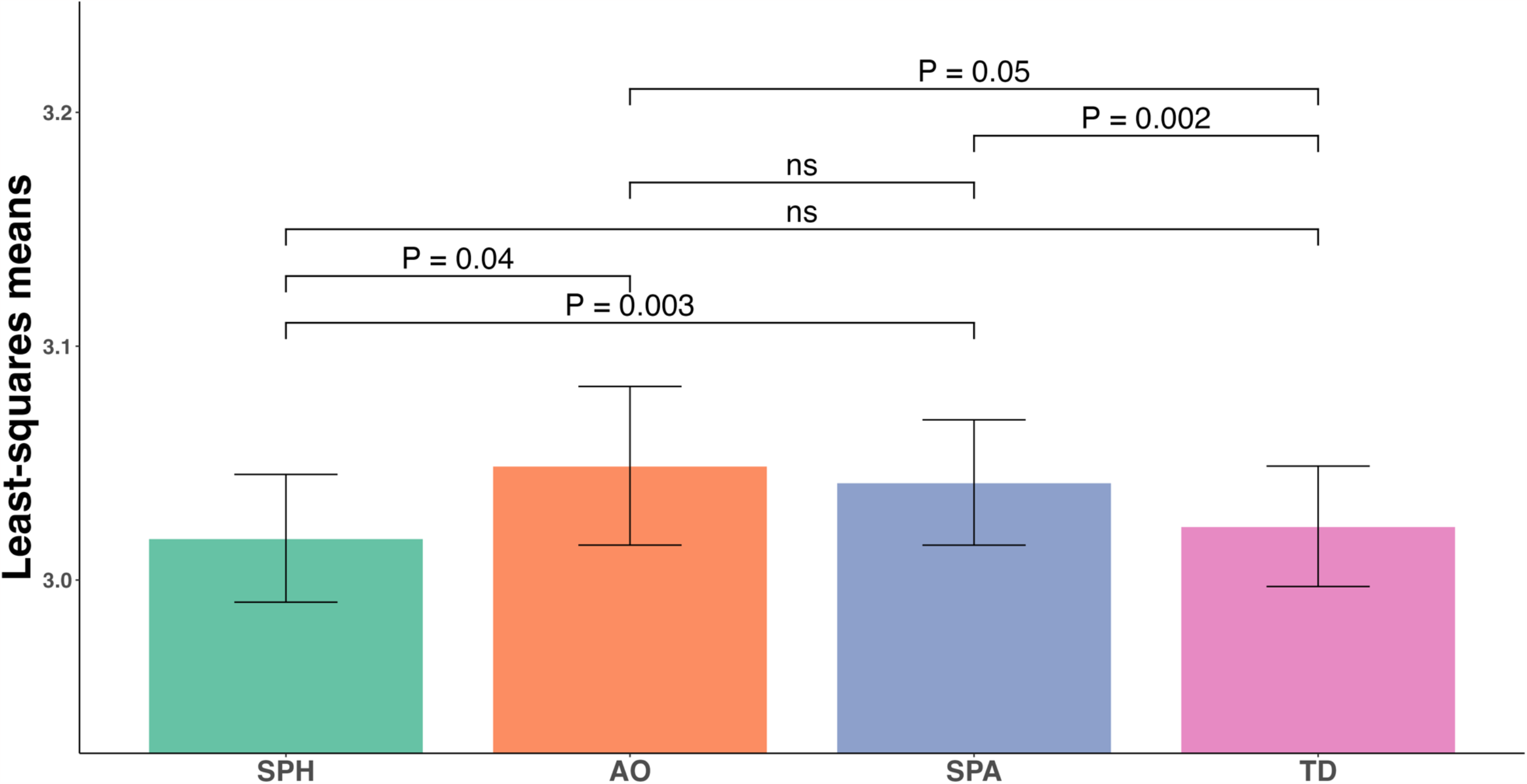
Bar plot demonstrating significant differences in thickness of left superior frontal gyrus in girls. Notably, the p values adjusted by the Benjamini-Hochberg procedure between each pair of comorbidity profiles are shown, while “ns” represents “non-significant”. Error bars represent the 95% confidence interval of the least-squares means for each profile.

### 3.4 Clinical correlates

Baseline analysis using one-way ANOVA across profiles and sex revealed significant differences in externalizing, internalizing and total problems scores on the CBCL, with higher scores indicating greater psychological deficits. Likewise, analyses comparing scores across all three profiles to typically developing children demonstrated significant differences (eTable 2).

For externalizing, internalizing, and total problems scores, post-hoc analyses demonstrated that all three profiles for both boys and girls were significantly higher than typically developing children (Figure 2). Both boys and girls in the AO group exhibited the most externalizing symptoms (boys: 22.45, 95% CI 21.21 - 23.70, P <0.001, girls: 22.73, 95% CI 20.95 - 24.51, P <0.001). Boys with the AOS profile had the most internalizing symptoms (22.55, 95% CI 20.41 - 24.69 P <0.001), while girls with the AO profile had the most internalizing symptoms (18.73, 95% CI 16.82 - 20.65, P <0.001). Similarly, the AOS profile and the AO profile had the highest total problem scores among boys (26.12, 95% CI 23.98 - 28.26, P<0.001) and girls (25.11, 95% CI 23.22 - 27.01, P <0.001), respectively.

**Figure 2.**
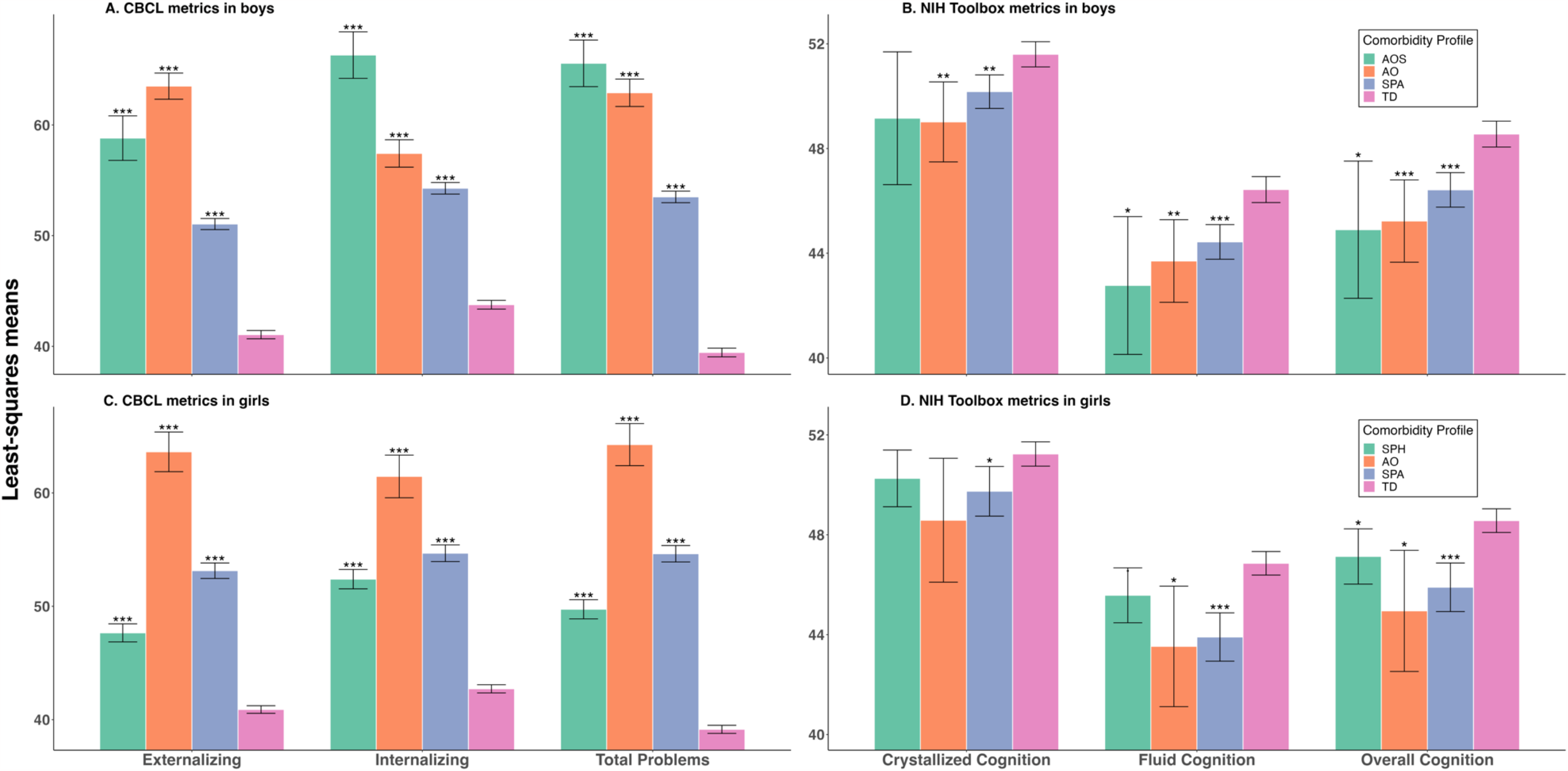
Bar plots demonstrating differences in CBCL metrics and NIH Toolbox metrics in boys and girls across all comorbidity profiles. Notably, the p values adjusted by Benjamini-Hochberg procedure for all three profiles compared to TD children. “***”, *P* ≤ 0.001; “**”, 0.001 < *P* ≤ 0.01; “*”, 0.01 < *P* ≤ 0.05; “.”, 0.05 < *P* ≤ 0.1; “”, 0.1 < *P*. Error bars represent 95% confidence interval of the least-squares means for each profile.

Post-hoc analyses using the NIH-Toolbox metrics revealed significantly lower total cognition scores for all three profiles in both boys and girls when compared to TD children, with the AOS profile having the lowest scores among boys (Estimate = -3.65, 95% CI -6.32 - -0.99, P = 0.01) and the AO profile having the lowest scores among girls (Estimate = - 3.61, 95% CI - 6.09 - -1.14, P = 0.01) (Figure 2). Crystallized cognition scores were lower among boys in the AO and SPA profiles, with the AO profile having the lowest score (Estimate = -2.58, 95% CI -4.18 - -0.98, P = 0.005), but not for those in the AOS profile when compared to typically developing children. Among girls, only those in the SPA profile demonstrated lower values compared to typically developing children (Estimate = -1.49, 95% CI -2.60 - -0.39, P = 0.05.) For fluid cognition, only girls in the SPA and AO groups had significantly lower values relative to TD children. In contrast, all three profiles for boys had significantly lower scores, with the AOS scores being the lowest among boys, (Estimate = -3.66, 95% CI -6.34 - -0.99, P = 0.01) and the AO group among girls (Estimate = -3.33, 95% CI -5.79 - -0.87, P = 0.02.)

## Discussion

In this study, we utilized novel psychiatric comorbidity network analysis to identify 3 distinct comorbidity profiles in youth with psychiatric disorders. Only boys showed greater than 50% prevalence of comorbid ADHD and OCD in the AOS profile, aligning with previous research indicating greater OCD symptoms at an earlier age often accompanied by ADHD, with girls having lower rates of comorbid OCD and ADHD.(34, 35) Boys, but not girls, with the AOS profile also showed a high rate of comorbid specific phobia and OCD, consistent with adult studies showing that men with anxiety disorders are more likely to have comorbid ADHD compared to women.(36) Studies suggest that women with specific phobia are more likely to have to have comorbid depression and anxiety disorders, with phobias often reported as preceding mood disorders that develop later in adolescence.(37, 38) The lack of such a profile in our analysis despite the presence of a specific phobia profile in girls may be due to lower prevalence of anxiety and depressive disorders within the ABCD cohort (Table 3) and the younger age of girls in this cohort. Mood/anxiety disorders typically emerge in mid to late adolescence.(39) With the average age of ABCD participants being 9-10 years old, comorbidity with mood and anxiety disorders in girls emerge over time.

We found that the cortical thickness of the superior frontal gyrus was marginally greater in girls across comorbidity profiles, but not in boys. The superior frontal gyrus is implicated in various psychiatric symptoms, such as response inhibition and introspection, and increased cortical thickness in this region has been linked with higher levels of irritability. (7, 40-42) The high prevalence of ADHD and ODD in both the AO and SPA profiles, including diagnoses known for their high levels of irritability and poor response inhibition, may indicate the presence of a sex-specific neurobiological mechanism for these comorbidity profile.(43-45) In adults, cortical thickness of the superior frontal gyrus has been shown to be greater in women than men, with data suggesting that structural sexual dimorphism may be related to changes in hormone receptor density differences.(46)

Our study also demonstrated key findings concerning behavioral profiles. Boys with the AOS profile exhibited the highest scores on the internalizing problems scale, while those with the AO profile showed the highest scores on the externalizing problems scale. Similarly, AOS boys and AO girls demonstrated the lowest total and fluid cognition scores, and SPA girls and AO boys had the lowest crystallized cognition scores. These findings align with the literature indicating that certain forms of ADHD tend to be more internalizing versus externalizing in some patients, as well as the known internalizing characteristics of OCD and specific phobia.(47) Cognitive deficits have been observed in ADHD, specifically crystallized and fluid cognition, consistent with our results for the NIH toolbox. Furthermore, given the large prevalence of ADHD in all profiles, our findings suggest a central role of inattention in cognitive deficits observed with comorbidity.(48, 49)

In contrast to previous findings and public perception, girls with the AO profile had higher levels of both internalizing and externalizing scores.(50) The fact that the female AO group displayed the highest internalizing and externalizing scores and lowest cognition scores stands in contrast to conventional wisdom, which posits that girls tend to exhibit more internalizing features, as well as fewer associated cognitive deficits in ADHD and mental illness more broadly.(51-53) This finding further highlights the potential role of data science in identifying groups of individuals with unique presentations. Employing a longitudinal approach would help strengthen these assumptions and determine the long-term impact these profiles could potentially have.

The primary limitations of our study stem from its cross-sectional design. Future research should examine how these comorbidity profiles evolve over time in relation to changing prevalence rates and neuromaturation. As with most data-driven approaches, interpretability remains a challenge as the model is currently unable to concretely identify the impact of each pair of comorbid diagnoses on whether a participant is placed in one profile versus another.(54) Nevertheless, the predominant comorbidities align remarkably well with existing literature, suggesting a potential role for data driven approaches in developing a more nuanced understanding of psychiatric disorders and their interconnectivity. Additionally, certain diagnoses were notably limited or absent within the ABCD cohort, including bipolar disorder, major depressive disorder, and DMDD; a common limitation of large-scale datasets.(55) We expect that the continued long-term follow up of the ABCD cohort will likely reveal a greater prevalence of the illnesses not observed in this study, as certain mood disorders are known to have prevalence changes across the life span, which we aim to explore in future studies.

In conclusion, our comorbidity network analyses successfully identify three comorbidity profiles across gender that aligned with existing literature on comorbidity, further validating the use of data-driven approaches in psychiatry. Further longitudinal analyses will be essential to shed light on the clinical impact of these comorbidity profiles, as well as the contribution of functional and structural connectivity in combination with the structural changes observed in this study.

## Data Availability

All data produced in the present study are available upon reasonable request to the authors

## Acknowledgements

Dr. Zhu is supported by NIH K01MH122774 and by a NARSAD Young Investigator Grant from the Brain & Behavior Research Foundation 27040. Dr. Luna is supported by the Moynihan Clinical Research Fellowship from Columbia University College of Physicians and Surgeons.

