## Supplemental data for "PSYCHIATRIC COMORBIDITY & STRUCTURAL BRAIN FEATURES IN THE ADOLESCENT BRAIN AND COGNITIVE DEVELOPMENT STUDY COHORT: A CROSS-SECTIONAL US POPULATION-BASED STUDY"

**eTable 1. Prevalence values for the top 10 dyads for each comorbidity profile among boys and girls.** Each profile was characterized by those comorbidity pairs that reached a threshold of greater than 50% prevalence within each profile. Notably, no pair reached the 50% threshold in the sparse comorbidity group for both boys and girls, and the specific phobia group in girls.

| **Prevalence, %** | | | | | | | | | | | |
|---|---|---|---|---|---|---|---|---|---|---|---|
| **Boys** | | | | | | **Girls** | | | | | |
| **AOS (n = 74)**[a] | | **AO (n = 214)** | | **SPA (n = 1200)** | | **SPH (n = 425)** | | **AO (n = 88)** | | **SPA (n = 577)** | |
| ADHD+SPH | 63.5 | ADHD+ODD | 73.8 | OCD+SPH | 4.3 | OCD+SPH | 9.7 | ADHD+ODD | 52.3 | ADHD+ODD | 5.4 |
| ADHD+OCD | 62.2 | ADHD+CD | 33.6 | ADHD+OCD | 4.3 | ADHD+SPH | 8.7 | ADHD+OCD | 34.1 | ADHD+OCD | 3.6 |
| OCD+SPH | 52.7 | CD+ODD | 30.8 | ADHD+SPH | 3.8 | | | ADHD+SPH | 29.6 | CD+ODD | 1.6 |
| ADHD+GAD | 36.5 | ADHD+OCD | 15.9 | ODD+SPH | 1.3 | | | OCD+ODD | 29.6 | CD+OCD | 1.4 |
| OCD+ODD | 35.1 | OCD+ODD | 15.0 | CD+OCD | 1.1 | | | CD+ODD | 26.1 | OCD+ODD | 1.0 |
| ADHD+ODD | 33.8 | ADHD+SPH | 14.0 | OCD+ODD | 0.8 | | | ODD+SPH | 25.0 | ADHD+CD | 0.5 |
| ODD+SPH | 31.1 | ODD+SPH | 13.6 | CD+SPH | 0.7 | | | ADHD+CD | 25.0 | | |
| GAD+OCD | 28.4 | CD+OCD | 10.8 | | | | | OCD+SPH | 18.2 | | |
| GAD+SPH | 27.0 | CD+SPH | 6.1 | | | | | CD+SPH | 17.1 | | |
| GAD+ODD | 23.0 | ADHD+SOC | 5.1 | | | | | GAD+SPH | 15.9 | | |

Abbreviations: **AOS**, ADHD+OCD+SPH; **AO**, ADHD+ODD; **SPH,** Specific Phobia; **SPA**, Sparse Clusters.

[a] This table only includes the present diagnoses with prevalence over 5% and the top 10 dyadic comorbidities.

**eTable 2. Chi-square analysis of top 4 diagnoses across all comorbidity profiles in boys and girls**

| | **Participants, No. (%)** | | | | | | | | | | | | | |
|---|---|---|---|---|---|---|---|---|---|---|---|---|---|---|
| | **Boys** | | | | | | | **Girls** | | | | | | |
| **Diagnoses** | **Overall** (N = 1488) | **AOS** (n = 74) | **AO** (n = 214) | **SPA** (n = 1200) | **Chi-squared value** | **Degrees of Freedom** | ***P* value** | **Overall** (N = 1090) | **SPH** (n = 425) | **AO** (n = 88) | **SPA** (n = 577) | **Chi-squared value** | **Degrees of Freedom** | ***P* value** |
| **ADHD** | 645 (43) | 62 (84) | 189 (88) | 394 (33) | 279.52 | 2 | <0.001 | 296 (27) | 37 (8.7) | 62 (70) | 197 (34) | 170.77 | 2 | <0.001 |
| **ODD** | 356 (24) | 33 (45) | 182 (85) | 141 (12) | 554.35 | 2 | <0.001 | 214 (20) | 19 (4.5) | 58 (66) | 137 (24) | 187.54 | 2 | <0.001 |
| **OCD** | 450 (30) | 55 (74) | 41 (19) | 354 (30) | 80.938 | 2 | <0.001 | 301 (28) | 41 (9.6) | 41 (47) | 219 (38) | 115.36 | 2 | <0.001 |
| **SPH** | 471 (32) | 54 (73) | 33 (15) | 384 (32) | 84.531 | 2 | <0.001 | 471 (43) | 425 (100) | 46 (52) | 0 (0) | 1000.5 | 2 | <0.001 |

Abbreviations: **AOS**, ADHD+OCD+SPH; **AO**, ADHD+ODD; **SPH**, Specific Phobia; **SPA**, Sparse Clusters.

**eTable 3. ANOVA analysis comparing CBCL between comorbidity profiles and typically developing children**

| | Boys | | | | | Girls | | | | |
|---|---|---|---|---|---|---|---|---|---|---|
| **CBCL** | **Sum of Squares** | **Degrees of Freedom** | **Mean Square** | **F value** | ***P* value** | **Sum of Squares** | **Degrees of Freedom** | **Mean Square** | **F value** | ***P* value** |
| **Externalizing** | | | | | | | | | | |
| **Between Groups** | 159926 | 3 | 53309 | 684.4 | <0.001 | 110361 | 3 | 36787 | 524.7 | <0.001 |
| **Within Groups** | 280164 | 3597 | 78 | | | 243407 | 3472 | 70 | | |
| **Total** | 440090 | 3600 | | | | 353768 | 3475 | | | |
| **Internalizing** | | | | | | | | | | |
| **Between Groups** | 128067 | 3 | 42689 | 502.0 | <0.001 | 107030 | 3 | 35677 | 440.6 | <0.001 |
| **Within Groups** | 305875 | 3597 | 85 | | | 281138 | 3472 | 81 | | |
| **Total** | 433942 | 3600 | | | | 388168 | 3475 | | | |
| **Total Problems** | | | | | | | | | | |
| **Between Groups** | 247796 | 3 | 82599 | 968.6 | <0.001 | 171160 | 3 | 57053 | 722.7 | <0.001 |
| **Within Groups** | 306746 | 3597 | 85 | | | 274108 | 3472 | 79 | | |
| **Total** | 554542 | 3600 | | | | 445268 | 3475 | | | |

**eFigure 1. Baseline diagnostic data analysis flowchart demonstrating pipeline for participant selection**

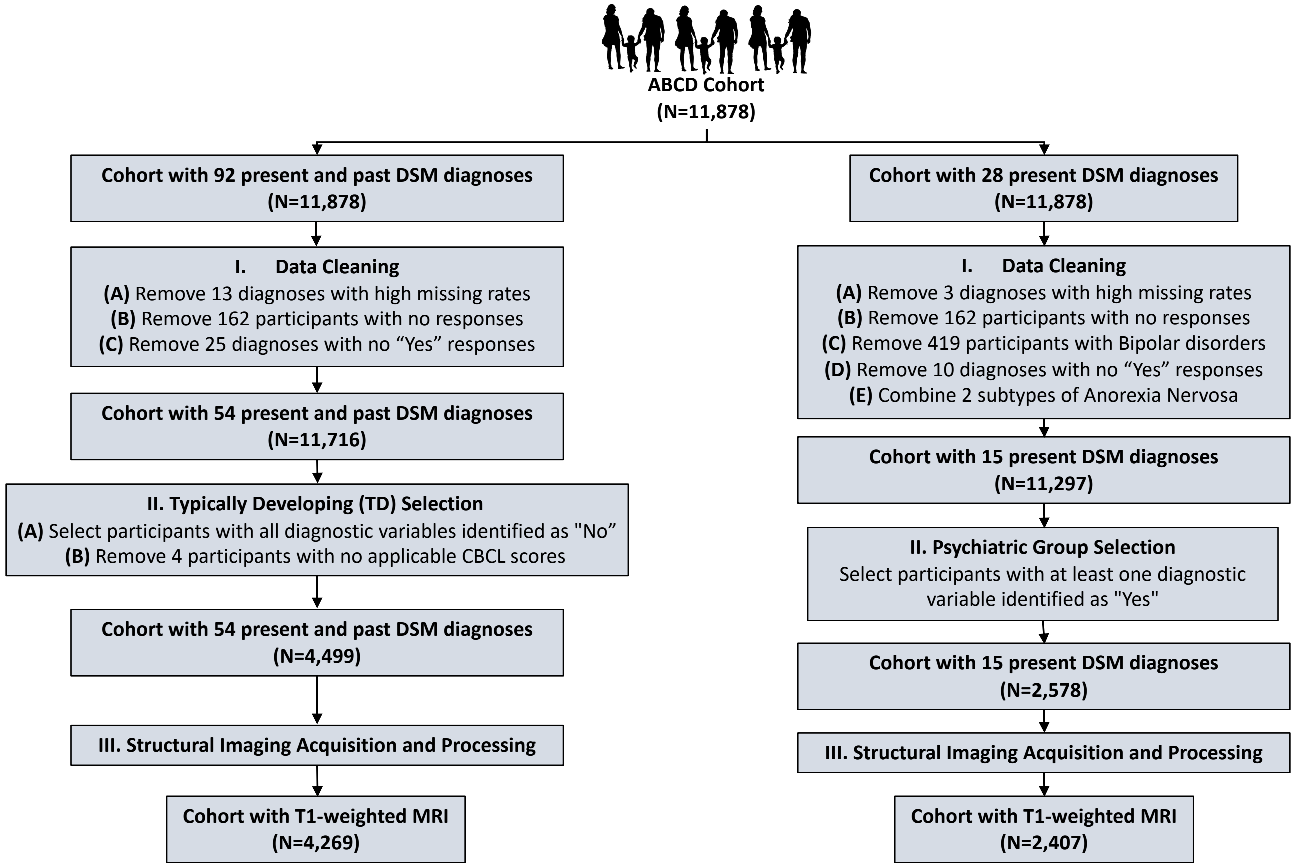

### Statistical Analysis

Data cleaning and all statistical analyses were performed in R. For LCA, we employed expectation-maximization implemented in the poLCA R package.[1] The demographic, clinical and behavioral characteristics were reported using mean with standard deviation, frequency with proportion as appropriate.

### Dyadic Cluster Analysis

In this study, we analyzed the frequencies of co-occurring diagnoses, referred to as "dyads", to determine diagnostic comorbidity networks. From a network perspective, comorbidity in psychiatry can be described as a simple undirected graph $G\ (V, E)$. The diagnoses are represented by a set of vertices $V$ with their co-occurrence represented by a set of edges $E$. This graph can be conveniently represented by an adjacency matrix, as shown in the dyadic comorbidity adjacency matrix in eFigure 2.

Specifically, for each subject $i$ $(1 \leq i \leq n; i \in N)$, a binary diagnosis vector $A_i = [A_{ij};\ j \in \{1, \dots, m\}]$ identifies whether the $i$ -th subject suffers from the $j$ -th diagnosis among $m$ diagnoses. From this vector, we define a dyadic comorbidity adjacency matrix of $i$-th subject, $W_i = \left\{W_{ijk};\ j, k \in \{1, \dots, m\}\right\}$, using the dyad (or outer product) of the $A_i$. In other words, $W_{ijk} = A_{ij}\ A_{ik}$ for $j, k \in \{1, \dots, m\}$. Consequently. the comorbidity pattern vector of $i$ -th subject, $X_i$, is formed by the elements from the diagonal and lower diagonal element of $W_i$. Thus, we can define $X_i = [W_{ijk};\ 1 \leq j \leq k \leq m]$.

For our current analysis, since we did not utilize specific attributes for each psychiatric diagnosis, elements of $A_i$ are binary, with 0 indicating the absence of the diagnosis and 1 indicating its presence. Consequently, the elements of $W_{ijk}$ also take binary values, with 0 denoting the absence of comorbidity between two diagnoses and 1 denoting the presence of the comorbidity. Conventionally, dyadic vectors do not include diagonal elements of a matrix (i.e., $j = k$). However, for our analysis, we extended the definitions to include diagonal elements to account for the presence of a psychiatric diagnosis without comorbidity.

Given the binary nature of the comorbidity data, traditional clustering methods such as K-Means, which rely on a Euclidean distance matrix, fail to identify meaningful clusters in this case. Alternatively, we employed Latent Class Analysis (LCA), a subset of structural equation modeling, to classify each individual into multiple mutually exclusive and exhaustive latent classes based on multivariate categorical data.

To incorporate the heterogeneity comorbidity patterns in the data, we assume there are $C$ conceptual populations of psychiatric disease clusters. Suppose also that independent random samples of clusters are drawn from the $C$ populations. Our primary interest is in making inferences about the unknown overall proportions of affected units in the $I$ populations, denoted by $p_1, p_2, .., p_C$, utilizing the distributions created by random sampling of clusters. Let $y_{ijk}$ be the probability of a positive response on the comorbidity between diagnosis $j$ and $k$ for an individual $i$ in category $c$ and let $p_c$ be the prior probability that a randomly chosen individual is in class $c$. Thus, the loglikelihood function is

$$f(p_1, p_2, .., p_C) = \sum_{c=1}^{C} \quad p_c \prod_{1 \le j \le k \le m} \quad y_{ijk}^{W_{ijk}} (1 - y_{ijk})^{1 - W_{ijk}}.$$

Then, the posterior probability that an individual with a comorbidity pattern vector $X_i$ belongs to category k is:

$$h(X_i) = \frac{p_c \prod_{1 \le j \le k \le m} \quad y_{ijk}^{W_{ijk}} (1 - y_{ijk})^{1 - W_{ijk}}}{\sum_{c=1}^{C} \quad p_c \prod_{1 \le j \le k \le m} \quad y_{ijk}^{W_{ijk}} (1 - y_{ijk})^{1 - W_{ijk}}}.$$

The parameters of the above model can be estimated via an EM algorithm. The optimal number of clusters is determined by Bayesian Information Criteria (BIC).[2]

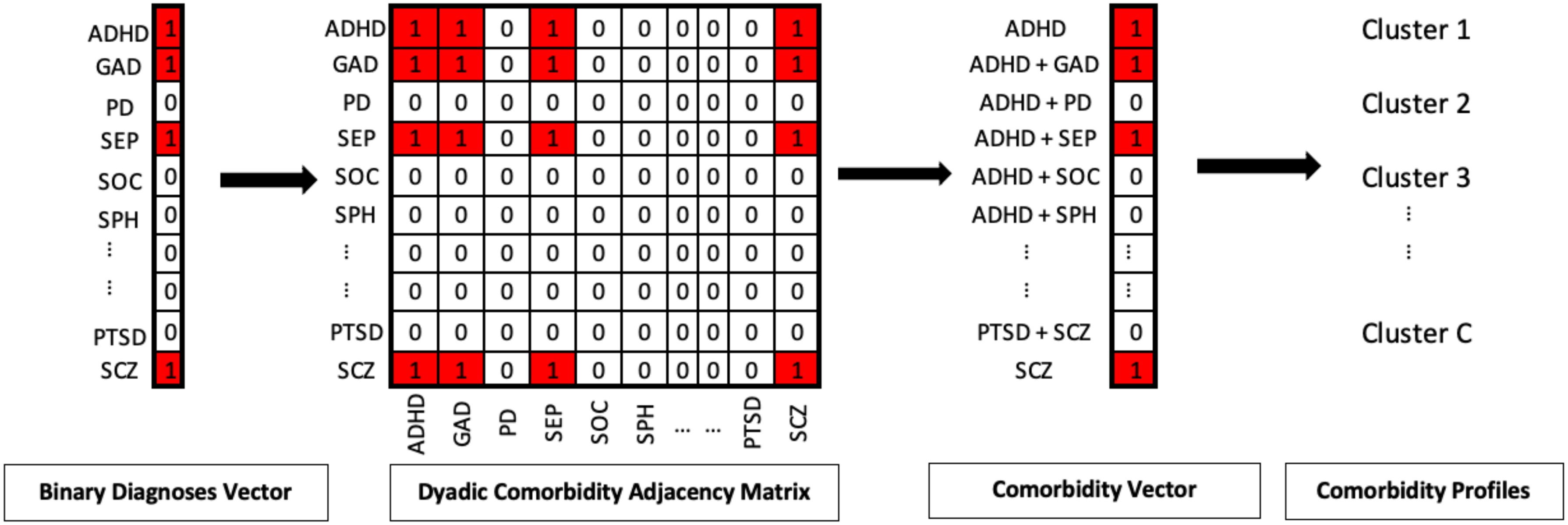


**eFigure 2. Depiction of dyadic profile analysis**

**eFigure 3. Network plots demonstrating comorbidity profiles across the AOS and AO profiles in boys and the AO profile in girls.** Darker lines indicate greater levels of comorbidity, and sphere sizes indicate prevalence level within the profile. Notably, each figure only includes the present diagnoses with prevalence over 5%. “External” refers to diagnosis considered to have typically externalizing features and “Internal” refers to diagnoses considered to have typically internalizing features.

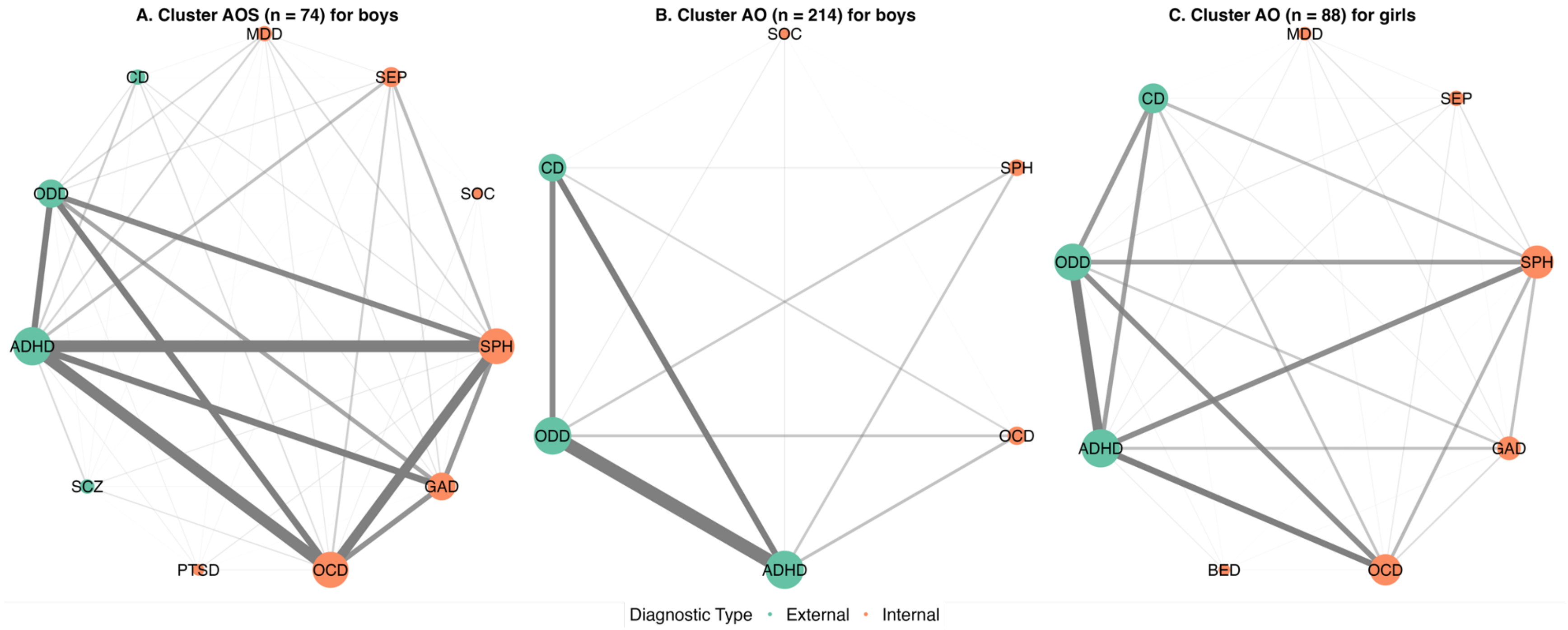

**eFigure 4. Bar plots demonstrating differences in other CBCL metrics in boys and girls across all comorbidity profiles.** Abbreviations: Anxdep, Anxious-Depressed; WithDep, Withdrawn-Depressed; Somatic, Somatic Complaints; Aggressive, Aggressive Behavior; RuleBreak, Rule-Breaking Behavior; Attention, Attention Problems; Social, Social Problems; Thought, Thought Problems. Error bars represent 95% confidence interval of the least-squares means for each profile.

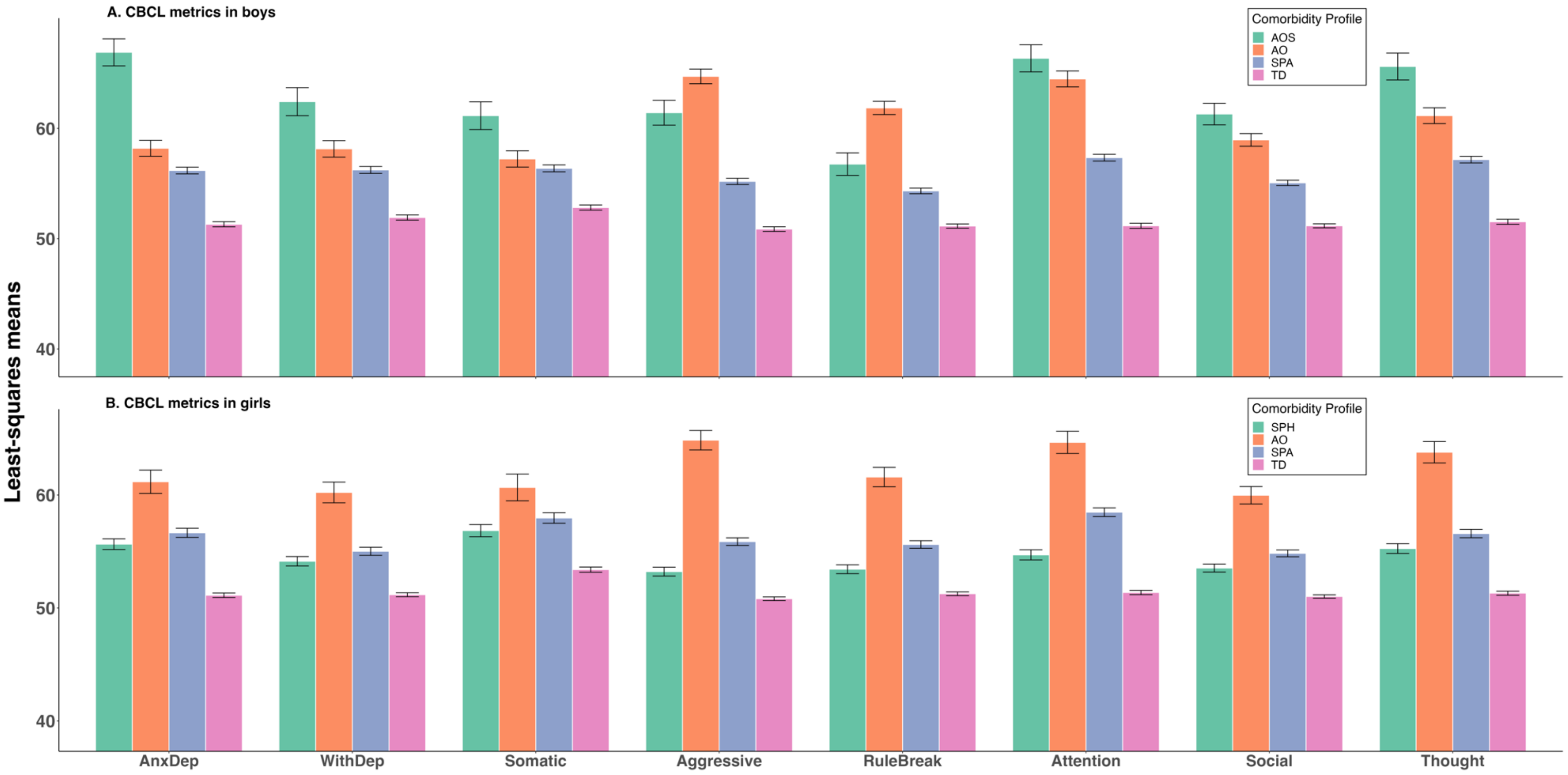
